# Procedure-Specific Long-Term Thromboembolic Risk Associated With Postoperative Atrial Fibrillation After Cardiac Surgery: A Systematic Review and Meta-Analysis

**DOI:** 10.64898/2026.08.23.26361121

**Authors:** Asad Ullah

## Abstract

Postoperative atrial fibrillation (POAF) is a frequent complication following cardiac surgery and has been associated with an increased risk of thromboembolic events. However, cardiac surgical populations are heterogeneous, and the long-term thromboembolic implications of POAF may differ according to the index surgical procedure. This systematic review and meta-analysis evaluated the procedure-specific association between POAF and long-term thromboembolic outcomes after adult cardiac surgery, with particular emphasis on coronary artery bypass grafting (CABG) and isolated valve surgery. PubMed and Scopus were searched from database inception through August 3, 2026. Studies reporting long-term thromboembolic outcomes in patients with new-onset POAF compared with patients without POAF were evaluated, with eligible evidence classified according to the index surgical procedure. Four observational studies were included in the primary quantitative synthesis, with two studies contributing to the CABG analysis and two to the isolated valve-surgery analysis. Adjusted hazard ratios (HRs) were pooled separately by procedure using inverse-variance methods, and a formal between-subgroup interaction test was performed. Following CABG, POAF was associated with an increased long-term thromboembolic hazard (pooled HR 1.147, 95% CI 1.053-1.249; I^2 =0%). A stronger association was observed following isolated valve surgery (pooled HR 1.362, 95% CI 1.181-1.573; I^2=0%). The between-subgroup interaction was statistically significant X^2 =(4.10, P=0.043), providing exploratory evidence that the magnitude of the association may differ according to surgical procedure. These findings suggest that the long-term thromboembolic implications of POAF may not be uniform across cardiac surgical populations. However, because only two studies contributed to each procedure subgroup and the available evidence was observational, the interaction should be considered hypothesis-generating. Further adequately powered studies with standardized outcome definitions and procedure-specific reporting are required to confirm these findings and determine their implications for long-term risk stratification and anticoagulation strategies.

## Introduction And Background

Postoperative atrial fibrillation (POAF) is one of the most common rhythm disturbances encountered after cardiac surgery and remains an important contributor to postoperative morbidity. Its development is multifactorial, involving interactions between a susceptible atrial substrate and perioperative factors such as inflammation, oxidative stress, autonomic disturbances, atrial ischemia, and changes in atrial loading conditions. Importantly, increasing evidence suggests that POAF should not always be considered an isolated and transient postoperative event because its clinical consequences may extend beyond the immediate perioperative period [1].

The mechanisms underlying POAF are complex and may vary according to patient characteristics and the type of cardiac operation performed. Structural atrial abnormalities, inflammatory responses, oxidative stress, neurohormonal activation, and electrophysiological changes occurring after cardiac surgery have all been implicated in its development. Recognition of these mechanisms has contributed to increasing interest in both prevention and treatment of POAF and in understanding its subsequent clinical consequences [2].

The longer-term importance of POAF has been reinforced by systematic evidence demonstrating associations between POAF and adverse outcomes following cardiac surgery. A systematic review and meta-analysis by Eikelboom et al. evaluated POAF after cardiac surgery and highlighted its association with clinically important postoperative outcomes. Such evidence challenges the traditional view that POAF can invariably be regarded as a benign, self-limited postoperative arrhythmia [3].

The potential thromboembolic consequences of POAF create an important management dilemma, particularly regarding anticoagulation. A systematic review by Yao et al. examining anticoagulation management of POAF after cardiac surgery demonstrated the uncertainty surrounding optimal anticoagulation strategies in this population. Decisions regarding anticoagulation must balance potential thromboembolic benefit against postoperative bleeding risk, while recognizing that evidence specific to POAF differs from that available for established nonsurgical atrial fibrillation [4].

Procedure-specific evidence is particularly important in patients undergoing coronary artery bypass grafting (CABG). Butt et al. examined the long-term thromboembolic risk associated with POAF following CABG and compared this population with patients with nonvalvular atrial fibrillation. Their findings contributed important evidence regarding the long-term thromboembolic significance of POAF after coronary surgery and demonstrated the need to evaluate POAF within the specific clinical context in which it occurs [5].

The association between POAF following CABG and subsequent cerebrovascular events has also been evaluated quantitatively. Megens et al., in a meta-analysis examining new-onset atrial fibrillation after CABG, specifically assessed its relationship with long-term stroke risk. Their findings further supported investigation of POAF as a potential marker of subsequent cerebrovascular risk rather than considering its relevance to be restricted to the index hospitalization [6].

Stroke after CABG, however, is multifactorial and cannot necessarily be attributed to POAF alone. Tarakji et al. examined the temporal onset, risk factors, and outcomes associated with stroke following CABG, demonstrating the importance of considering when cerebrovascular events occur and the multiple perioperative and patient-related factors contributing to stroke after coronary surgery [7].

Underlying patient characteristics further influence cerebrovascular risk following CABG. Mérie et al. evaluated stroke risk after CABG in relation to age and comorbidities, emphasizing that baseline clinical characteristics contribute substantially to postoperative and subsequent stroke risk. Consequently, any observed association between POAF and thromboembolic events must be interpreted within the broader vascular-risk profile of cardiac surgical patients [8].

Another important consideration is recurrence of atrial fibrillation after apparently transient POAF. Lowres et al. systematically reviewed the incidence of POAF recurrence among patients discharged in sinus rhythm after cardiac surgery and demonstrated that recurrent atrial fibrillation may occur following discharge. This observation provides a potential explanation for why an arrhythmia initially detected during the perioperative period may remain relevant during longer-term follow-up [9].

Given these uncertainties and the expanding observational literature, systematic assessment of the available evidence is warranted. The methodology and reporting of the present systematic review and meta-analysis were guided by the Preferred Reporting Items for Systematic Reviews and Meta-Analyses (PRISMA) 2020 recommendations [10].

The methodological quality of eligible nonrandomized studies was assessed using the Newcastle-Ottawa Scale (NOS), which evaluates study selection, comparability of groups, and ascertainment of outcomes or exposures in observational research [11].

Because long-term thromboembolic outcomes are frequently reported as time-to-event estimates, appropriate methods are required for incorporating hazard ratios and related summary statistics into quantitative synthesis. The methodological principles described by Tierney et al. provide established approaches for incorporating summary time-to-event information into meta-analysis and informed the handling of such effect estimates in the present review [12].

More recent population-level evidence has further clarified the association between new-onset atrial fibrillation following coronary surgery and subsequent cerebrovascular risk. Taha et al. evaluated new-onset atrial fibrillation after coronary surgery in a nationwide cohort and specifically examined subsequent stroke risk, providing contemporary procedure-specific evidence relevant to the long-term consequences of POAF after CABG [13].

The clinical interpretation of these associations is particularly important because an observed increase in thromboembolic risk does not automatically establish an indication for indefinite anticoagulation.

Contemporary atrial fibrillation guidance emphasizes individualized assessment of thromboembolic and bleeding risks when determining antithrombotic management [14].

Additional long-term evidence has emerged from the CORONARY trial population. Conen et al. examined new-onset perioperative atrial fibrillation following CABG and its association with long-term adverse events, providing further evidence that the prognostic relevance of perioperative atrial fibrillation may extend beyond the immediate postoperative period [15].

Finally, broader systematic evidence continues to demonstrate the clinical importance of atrial fibrillation following cardiac surgery. The systematic review and meta-analysis by Caldonazo et al. evaluated AF after cardiac surgery and its association with adverse outcomes, reinforcing the need for continued investigation of the longer-term significance of this common postoperative complication [16].

Despite this growing literature, an important question remains unresolved: whether the magnitude of long-term thromboembolic risk associated with POAF differs according to the cardiac surgical procedure performed. Combining CABG and valve-surgery populations may obscure clinically relevant differences in baseline risk, atrial substrate, postoperative management, and thromboembolic outcomes. The present systematic review and meta-analysis therefore evaluates the long-term thromboembolic association of POAF separately after CABG and isolated valve surgery and examines whether the magnitude of this association differs between the two surgical populations.

## Review

### Methods

This systematic review and meta-analysis was designed to evaluate the association between postoperative atrial fibrillation (POAF) and long-term thromboembolic outcomes after cardiac surgery, with particular emphasis on whether the magnitude of this association differs between patients undergoing coronary artery bypass grafting (CABG) and those undergoing isolated valve surgery.

### Reporting Framework

The methodology and reporting of this review were guided by the Preferred Reporting Items for Systematic Reviews and Meta-Analyses (PRISMA) 2020 recommendations [10]. The review question, eligibility criteria, study-selection process, data extraction, risk-of-bias assessment, and quantitative synthesis were defined before interpretation of the pooled results.

### Search Strategy and Information Sources

A systematic literature search was conducted in PubMed and Scopus from database inception through August 3, 2026, to identify studies evaluating long-term thromboembolic outcomes associated with new-onset POAF after adult cardiac surgery. The search strategy incorporated terms related to POAF, cardiac surgical procedures, and thromboembolic outcomes, including “postoperative atrial fibrillation,” “post-operative atrial fibrillation,” “new-onset atrial fibrillation,” “POAF,” “cardiac surgery,” “coronary artery bypass,” “CABG,” “valve surgery,” “valvular surgery,” “aortic valve replacement,” “mitral valve surgery,” “stroke,” “ischemic stroke,” “ischaemic stroke,” “thromboembolism,” “thromboembolic,” “embolism,” “transient ischemic attack,” “transient ischaemic attack,” and “TIA.” Search terms were combined using the Boolean operators AND and OR. Reference lists of relevant reviews and eligible primary studies were additionally examined to identify potentially eligible studies.The search concepts were combined as follows: (“postoperative atrial fibrillation” OR “post-operative atrial fibrillation” OR “new-onset atrial fibrillation” OR “POAF”) AND (“cardiac surgery” OR “coronary artery bypass” OR “CABG” OR “valve surgery” OR “valvular surgery” OR “aortic valve replacement” OR “mitral valve surgery”) AND (“stroke” OR “ischemic stroke” OR “ischaemic stroke” OR “thromboembolism” OR “thromboembolic” OR “embolism” OR “transient ischemic attack” OR “transient ischaemic attack” OR “TIA”).

### Eligibility Criteria

Studies were considered eligible when they evaluated adult patients undergoing cardiac surgery and assessed postoperative or new-onset atrial fibrillation as the exposure. For the principal procedure-specific analysis, studies were required to provide separately extractable data for CABG or isolated valve surgery.

The outcome of interest was long-term thromboembolism following POAF. Studies reporting adjusted time-to-event estimates, particularly hazard ratios (HRs) with corresponding 95% confidence intervals (CIs), were prioritized for quantitative synthesis. Studies were required to provide an appropriate comparison between patients who developed POAF and patients undergoing the same surgical procedure who did not develop POAF.

Studies were excluded from the primary quantitative analysis when procedure-specific data could not be separated, when POAF was not evaluated as the exposure of interest, when only immediate perioperative outcomes were reported without relevant longer-term follow-up, or when an appropriate POAF-versus-no-POAF effect estimate could not be extracted.

### Study Selection

Potentially relevant studies identified through the literature search were evaluated according to the predefined eligibility criteria. Studies were considered eligible when they evaluated adults undergoing cardiac surgery, assessed new-onset POAF as the exposure, included an appropriate comparison group without POAF, and reported long-term thromboembolic outcomes, including ischemic stroke, transient ischemic attack, systemic or peripheral embolism, or a defined thromboembolic composite. Studies were classified according to the index surgical procedure, with CABG and isolated valve-surgery populations evaluated separately when procedure-specific data were available.

Systematic reviews and meta-analyses were used to contextualize the existing evidence but were not treated as primary cohorts in the quantitative synthesis [3,4,6,9,16]. Primary studies providing relevant CABG evidence included Butt et al. [5], Taha et al. [13], and Conen et al. [15], although inclusion in individual quantitative analyses depended on compatibility of the reported endpoint. The primary procedure-specific quantitative synthesis incorporated compatible adjusted estimates from Butt et al. [5] and Taha et al. [13] for CABG and Butt et al. [17] and Rezk et al. [18] for isolated valve surgery.

### Data Extraction

Data were extracted according to the index cardiac procedure and included study characteristics, population size, number of patients developing POAF, comparison-group size, duration of follow-up, thromboembolic endpoint, effect measure, and corresponding 95% CI when available.

Where reported, information concerning the definition of POAF, anticoagulation use, adjustment variables, and outcome ascertainment was also considered because these factors could influence the relationship between POAF and subsequent thromboembolic events.

For CABG, the primary thromboembolism analysis incorporated compatible adjusted estimates from Butt et al. [5] and Taha et al. [13]. Conen et al. provided additional CABG evidence regarding long-term outcomes after new-onset perioperative atrial fibrillation [15]; however, its stroke-specific endpoint was distinguished from the broader thromboembolic composite used for the principal analysis.

### Outcome Definition

The principal outcome was long-term thromboembolism after development of POAF. Because thromboembolic outcome definitions were not completely identical among the included studies, individual study definitions were retained rather than assuming that every component of the composite endpoint was identical across cohorts.

Stroke-specific outcomes were distinguished from broader composite thromboembolic outcomes when necessary. This distinction was particularly important for the CABG evidence because the CORONARY analysis by Conen et al. reported long-term stroke-related outcomes [15], whereas the principal pooled analysis was based on studies reporting sufficiently comparable thromboembolic outcomes.

### Risk of Bias Assessment

The methodological quality of included observational studies was assessed using the Newcastle-Ottawa Scale (NOS) [11]. Assessment considered selection of the study population, comparability between POAF and non-POAF groups, ascertainment of outcomes, and adequacy of follow-up.

Particular attention was given to adjustment for clinically relevant factors that could influence long-term thromboembolic risk, including age, cardiovascular comorbidity, previous cerebrovascular disease, and anticoagulation when reported.

### Statistical Analysis

Adjusted HRs were preferentially used because the principal outcome was a long-term time-to-event endpoint. HRs and their corresponding 95% CIs were transformed to the logarithmic scale for quantitative synthesis.

Standard errors of the log HRs were derived from the reported 95% CIs using established methods for incorporating summary time-to-event information into meta-analysis [12]:

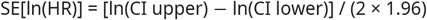

Inverse-variance weighting was subsequently applied to the log-transformed effect estimates.

CABG and isolated valve surgery were pooled separately rather than combining all cardiac operations into a single overall estimate. This procedure-specific approach was chosen because differences in underlying disease, atrial substrate, baseline vascular risk, and postoperative management could potentially modify the association between POAF and subsequent thromboembolism.

A random-effects framework was used for the principal procedure-specific analyses. Statistical heterogeneity was assessed using Cochran’s Q and the I^2 statistic. Because only two studies contributed to each principal procedure subgroup, estimates of I^2 and between-study variance were interpreted cautiously; absence of detected statistical heterogeneity was not considered evidence of complete clinical or methodological homogeneity.

A fixed-effect inverse-variance analysis was also examined as a sensitivity analysis.

### Procedure-Specific Interaction

After obtaining separate pooled estimates for CABG and isolated valve surgery, a formal between-subgroup interaction test was performed to evaluate whether the association between POAF and long-term thromboembolism differed according to surgical procedure.

The difference between the pooled log HRs was evaluated relative to its standard error. The resulting test statistic was used to calculate a two-sided P value for interaction. A P value <0.05 was considered statistically significant.

Importantly, statistical significance within one surgical subgroup and nonsignificance within another were not considered sufficient evidence of a procedure-specific difference. Interpretation of effect modification was based on the formal between-subgroup interaction test.

### Methodological Caution

The quantitative analysis was based on aggregate adjusted estimates rather than individual patient data. Differences in POAF ascertainment, thromboembolic endpoint definitions, follow-up, adjustment models, and anticoagulation practices were therefore considered when interpreting the pooled results.

Furthermore, because only two studies contributed to each principal procedure-specific pool, the meta-analysis was considered exploratory with respect to effect modification. Particular caution was applied to the interpretation of heterogeneity statistics and the procedure interaction.

## Results

### Study Selection

The literature search identified 247 records. After removal of 66 duplicate records, 181 unique records underwent title and abstract screening. Of these, 167 records were excluded, leaving 14 reports for full-text assessment. Following application of the predefined eligibility criteria, 10 full-text reports were excluded, and four studies were included in the primary quantitative synthesis [5,13,17,18]. Reasons for full-text exclusion included secondary/non-primary research, lack of separately extractable procedure-specific data, absence of POAF as the exposure of interest, absence of an appropriate long-term thromboembolic outcome or follow-up, and lack of an extractable POAF-versus-no-POAF effect estimate suitable for the primary quantitative analysis. Studies contributing to the quantitative synthesis were classified according to the index surgical procedure, with two studies contributing to the CABG analysis [5,13] and two to the isolated valve-surgery analysis [17,18].The study-selection process is summarized in the PRISMA 2020 flow diagram (Figure *1*).

**FIGURE 1:**
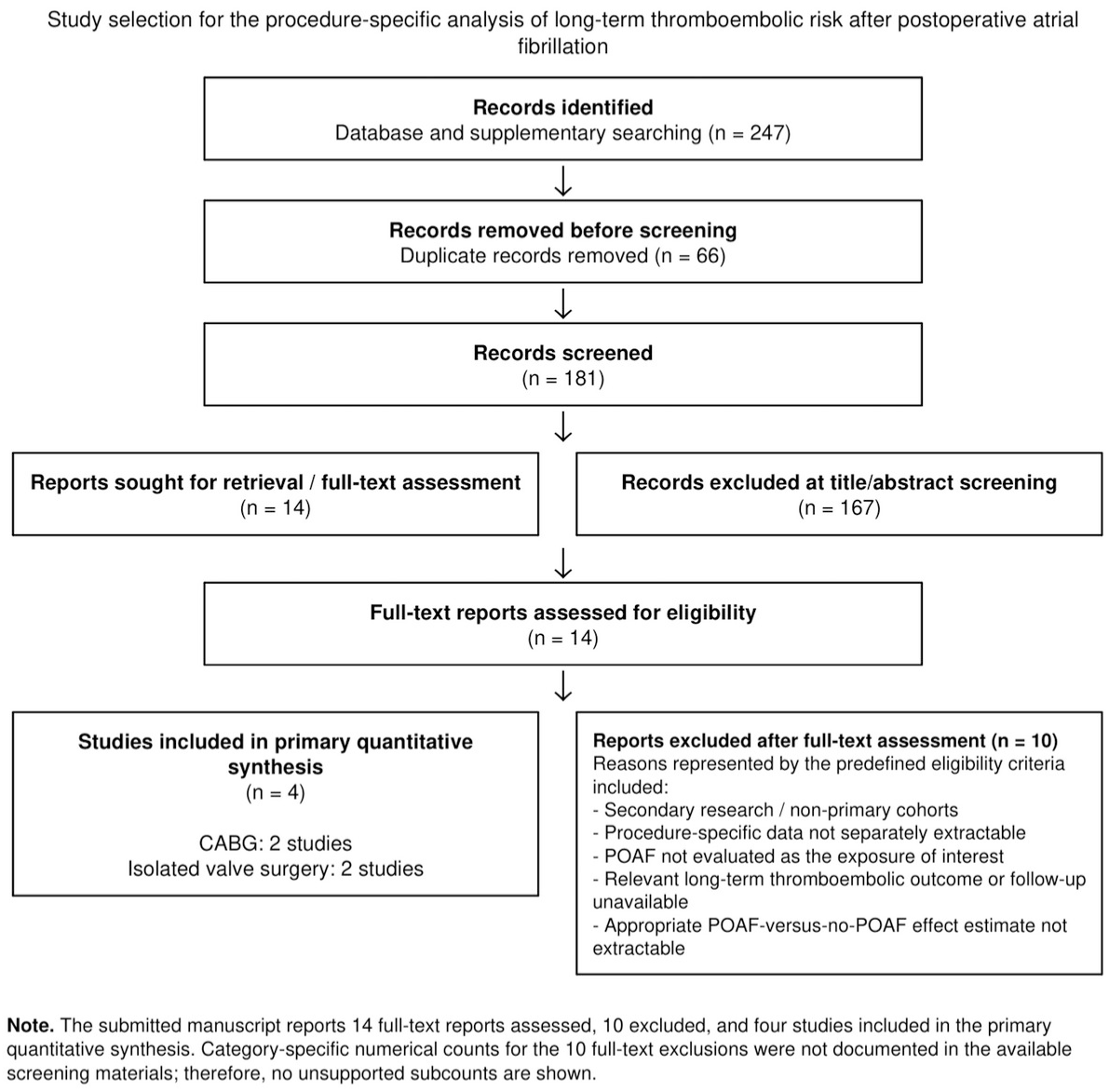
PRISMA 2020 flow diagram

Detailed study characteristics are presented for the four studies included in the primary quantitative synthesis. Full-text reports that did not meet the prespecified eligibility criteria were not included in the study-characteristics table because they did not contribute eligible data to the quantitative synthesis; their reasons for exclusion are summarized in the study-selection process and PRISMA flow diagram.

### Study Characteristics

Four observational studies met the prespecified criteria for inclusion in the primary quantitative synthesis [5,13,17,18]. Two studies evaluated patients undergoing coronary artery bypass grafting (CABG) [5,13], while two evaluated patients undergoing isolated valve surgery [17,18]. All four studies compared patients who developed postoperative atrial fibrillation (POAF) with corresponding surgical populations without POAF and reported adjusted time-to-event estimates suitable for the procedure-specific quantitative synthesis. Detailed characteristics of the included studies, including surgical procedure, population size, POAF and non-POAF groups, duration of follow-up, thromboembolic outcome, and adjusted effect estimates, are summarized in Table *3*.

**TABLE 1:** PICO framework used to define the review question and eligibility criteria. PICO, Population, Intervention/Exposure, Comparator, Outcome; POAF, postoperative atrial fibrillation; CABG, coronary artery bypass grafting; HR, hazard ratio; CI, confidence interval.

| Component | Definition used in this review |
| --- | --- |
| Population (P) | Adults undergoing cardiac surgery, with procedure-specific analyses for CABG and isolated valve surgery |
| Exposure (I) | New-onset postoperative atrial fibrillation (POAF) following cardiac surgery |
| Comparator (C) | Patients undergoing the same surgical procedure who did not develop POAF |
| Outcome (O) | Long-term thromboembolic outcomes, including ischemic stroke, transient ischemic attack, systemic/peripheral embolism, or a defined thromboembolic composite |
| Effect measure for quantitative synthesis | Adjusted time-to-event estimates, preferentially hazard ratios (HRs) with 95% confidence intervals |

**TABLE 2:** Literature search strategy used to identify studies evaluating long-term thromboembolic outcomes associated with postoperative atrial fibrillation after cardiac surgery. POAF, postoperative atrial fibrillation; CABG, coronary artery bypass grafting; TIA, transient ischemic attack.

| Search component | Search terms |
| --- | --- |
| Databases | PubMed and Scopus |
| Search period | Database inception through August 3, 2026 |
| POAF concept | "postoperative atrial fibrillation" OR "post-operative atrial fibrillation" OR "new-onset atrial fibrillation" OR "POAF" |
| Surgical procedure concept | "cardiac surgery" OR "coronary artery bypass" OR "CABG" OR "valve surgery" OR "valvular surgery" OR "aortic valve replacement" OR "mitral valve surgery" |
| Thromboembolic outcome concept | "stroke" OR "ischemic stroke" OR "ischaemic stroke" OR "thromboembolism" OR "thromboembolic" OR "embolism" OR "transient ischemic attack" OR "transient ischaemic attack" OR "TIA" |
| Combined search | (POAF terms) AND (cardiac-surgery terms) AND (thromboembolic-outcome terms) |
| Supplementary search | Reference lists of relevant reviews and eligible primary studies were examined for additional potentially eligible studies |

**TABLE 3:**
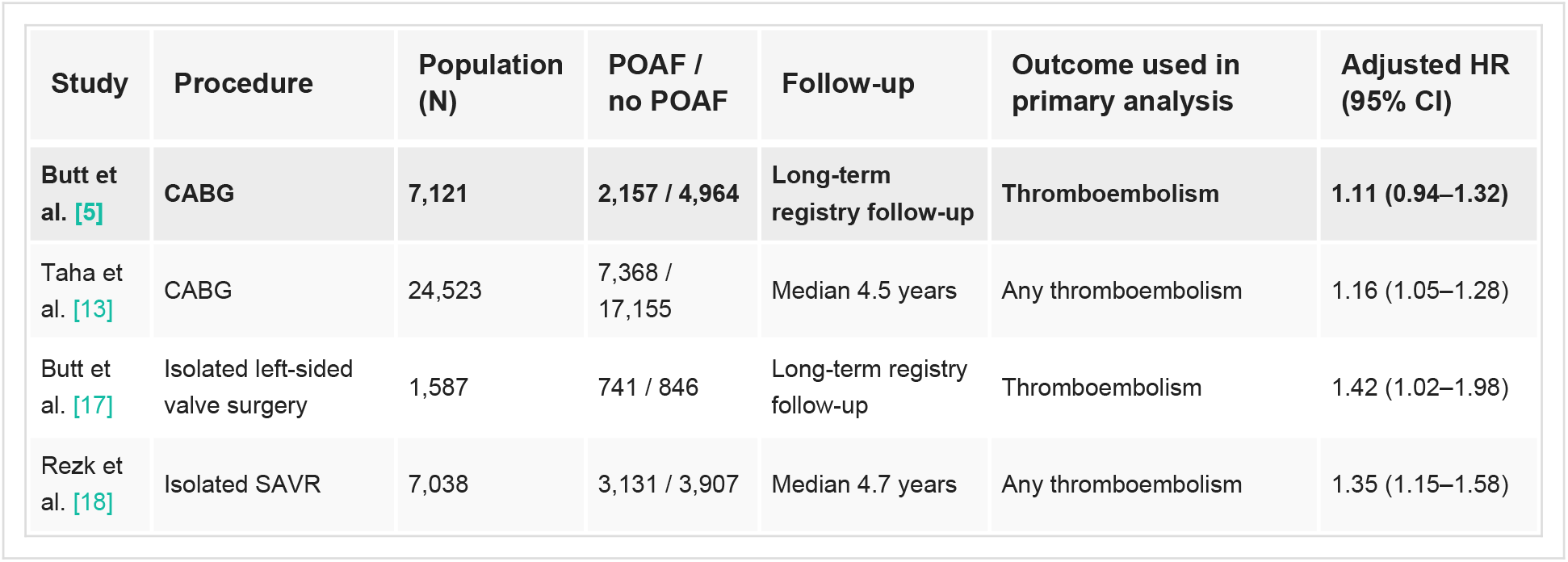
Detailed characteristics of the four studies included in the primary quantitative synthesis.

**TABLE 4:** Procedure-specific pooled estimates of long-term thromboembolic risk associated with postoperative atrial fibrillation.

| Surgical procedure | Studies | Total N | Pooled HR | 95% CI | I<sup>2</sup> |
| --- | --- | --- | --- | --- | --- |
| CABG [5,13] | 2 | 31,644 | 1.147 | 1.053-1.249 | 0% |
| Isolated valve surgery [17,18] | 2 | 8,625 | 1.362 | 1.181-1.573 | 0% |

### Risk of Bias

Risk of bias was assessed for the four observational studies included in the primary quantitative synthesis using the Newcastle-Ottawa Scale (NOS), considering cohort selection, comparability of the POAF and non-POAF groups, outcome ascertainment, and adequacy of follow-up. All included studies used observational cohort designs and adjusted analyses; nevertheless, residual confounding, differences in POAF ascertainment, anticoagulation exposure, outcome definitions, and follow-up remained potential sources of bias. These limitations were considered when interpreting the pooled estimates.

### Procedure-Specific Pooled Analysis

The individual study estimates and procedure-specific pooled estimates are presented in Figure *2*.

**FIGURE 2:**
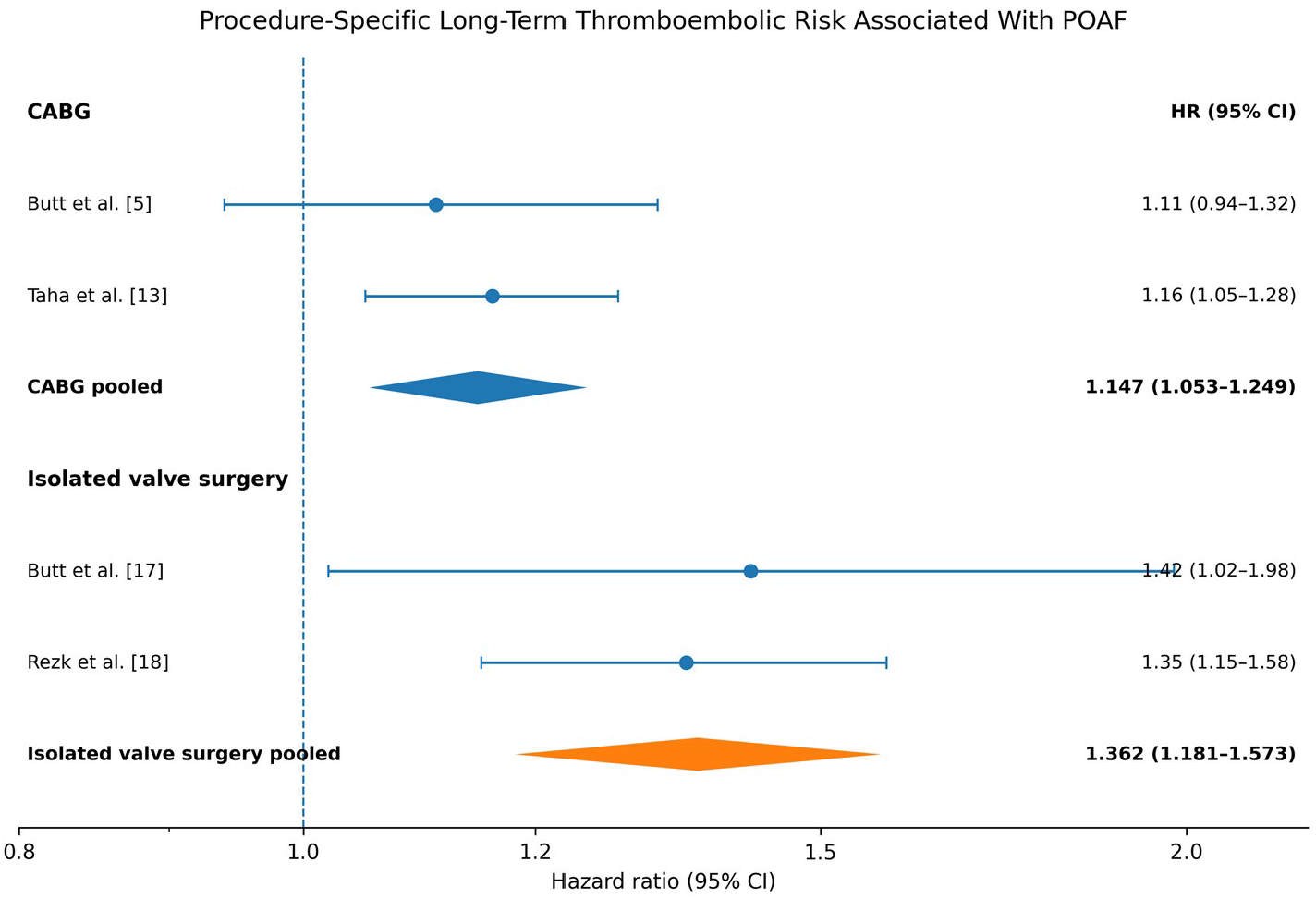
Forest plot of procedure-specific long-term thromboembolic risk associated with postoperative atrial fibrillation.

### Procedure-Specific Interaction Analysis

The formal between-subgroup interaction test yielded:

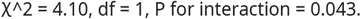

Thus, the difference between the CABG and valve-surgery pooled estimates reached conventional statistical significance. The association between POAF and subsequent thromboembolism was greater in the isolated valve-surgery subgroup than in the CABG subgroup.

Importantly, this interaction concerns the relative association between POAF and long-term thromboembolism within each surgical population. It should not be interpreted as demonstrating that valve-surgery patients necessarily have a higher absolute thromboembolic event rate than CABG patients.

### Fixed-Effect Sensitivity Analysis

Because the estimated between-study variance was zero within both procedure groups, the fixed-effect inverse-variance sensitivity analysis produced essentially the same estimates as the primary analysis.

For CABG, the pooled estimate remained HR 1.147 (95% CI 1.053-1.249) [5,13]. For isolated valve surgery, the corresponding estimate remained HR 1.362 (95% CI 1.181-1.573) [17,18]. The procedure-specific interaction remained P = 0.043.

The sensitivity analysis therefore did not materially alter the direction or magnitude of the principal findings.

### Summary of Principal Findings

Across the four studies included in the primary quantitative synthesis [5,13,17,18], POAF was associated with increased long-term thromboembolic risk after both CABG and isolated valve surgery. The magnitude of the association was smaller following CABG (HR 1.15) than following isolated valve surgery (HR 1.36), and formal testing provided evidence of a procedure-specific difference (P for interaction=0.043).

Because only two studies were available within each procedure subgroup, these results should be considered evidence of potential procedure-specific effect modification rather than definitive proof of a biological difference between CABG and valve surgery.

## Discussion

The present meta-analysis suggests that the long-term thromboembolic implications of postoperative atrial fibrillation (POAF) may differ according to the type of cardiac surgical procedure. POAF has traditionally been regarded as a predominantly transient postoperative arrhythmia arising from perioperative inflammation, autonomic disturbance, atrial injury, and hemodynamic stress [1,2]. However, accumulating evidence indicates that POAF may identify patients with persistent susceptibility to atrial fibrillation and subsequent thromboembolic events beyond the immediate postoperative period [3,4,6,9,16].

In the present procedure-specific analysis, POAF was associated with an increased hazard of long-term thromboembolism after both CABG and isolated valve surgery. The pooled relative association was smaller after CABG, with a pooled HR of 1.147 (95% CI 1.053-1.249) [5,13], than after isolated valve surgery, for which the pooled HR was 1.362 (95% CI 1.181-1.573) [17,18]. The formal subgroup interaction was statistically significant (χ^2 = 4.10, P = 0.043), providing exploratory evidence that the long-term thromboembolic implications of POAF may be procedure-specific.

### POAF After CABG

The CABG-specific findings were derived from two large cohorts [5,13]. Butt et al. reported an adjusted HR of 1.11 (95% CI 0.94-1.32) [5], whereas Taha et al. reported an adjusted HR of 1.16 (95% CI 1.05-1.28) [13]. Their pooled estimate of 1.147 indicates a modest but statistically significant increase in long-term thromboembolic hazard associated with POAF.

Several mechanisms could potentially explain this association. POAF may represent more than an isolated response to surgery and could instead identify patients with an underlying atrial substrate that predisposes them to recurrent atrial fibrillation. Evidence of recurrent AF following apparently transient postoperative episodes has been demonstrated in previous systematic evidence [9]. Consequently, some thromboembolic events occurring during longer follow-up may be related to recurrent or subsequently recognized AF rather than the original postoperative episode itself.

Nevertheless, POAF may also function as a risk marker rather than a direct causal factor. Patients who develop POAF frequently have characteristics that are themselves associated with adverse cardiovascular outcomes. Therefore, despite the use of adjusted estimates in the included cohorts, residual confounding cannot be excluded. The observed association should not automatically be interpreted as evidence that POAF itself causes subsequent thromboembolism.

The long-term thromboembolic consequences of POAF after CABG are clinically relevant because previous observational evidence has demonstrated concern regarding later thromboembolic outcomes in this population [5,6,13]. At the same time, the magnitude of risk associated with postoperative AF should not simply be assumed to be equivalent to that associated with established nonsurgical atrial fibrillation. This distinction is particularly important when considering long-term anticoagulation.

### POAF After Valve Surgery

A larger pooled relative association was observed among patients undergoing isolated valve surgery compared with CABG. The two valve-surgery studies yielded adjusted HRs of 1.42 (95% CI 1.02-1.98) [17] and 1.35 (95% CI 1.15-1.58) [18], resulting in a pooled HR of 1.362 (95% CI 1.181-1.573).

The greater relative association observed following valve surgery is biologically plausible, although the present analysis cannot establish its mechanism. Valve disease may coexist with structural atrial remodeling, atrial enlargement, pressure or volume overload, and chronic changes in atrial electrophysiology. In this setting, postoperative AF could potentially represent the clinical manifestation of a more established atrial substrate rather than a purely transient postoperative phenomenon.

However, this interpretation should remain cautious. The significant interaction observed in the present analysis demonstrates a statistical difference between the pooled relative effect estimates; it does not demonstrate why that difference exists. Furthermore, valve surgery represents a heterogeneous clinical category. Aortic and mitral valve disease differ substantially in their effects on atrial structure, hemodynamics, baseline AF susceptibility, and thromboembolic risk. The present evidence base is too limited to establish whether the observed association is consistent across individual valve procedures.

### Procedure-Specific Effect Modification

The principal finding of this analysis is evidence of an interaction between surgical procedure and the association of POAF with long-term thromboembolism, with the formal interaction test reaching conventional statistical significance (P=0.043). The pooled HR was approximately 1.15 after CABG compared with 1.36 after isolated valve surgery, with P for interaction = 0.043.

This result is important because statistical significance within one subgroup and nonsignificance within another does not, by itself, establish that the subgroup effects differ. A formal interaction test is required. In the present analysis, direct comparison of the subgroup estimates provided evidence for a difference.

Nevertheless, the interaction should be considered exploratory. Only two studies contributed to each procedure subgroup. Consequently, the interaction estimate is dependent on a small number of study-level estimates, and its P value lies close to the conventional 0.05 threshold. The finding therefore requires confirmation in additional independent cohorts.

The results also should not be interpreted as demonstrating that valve-surgery patients have a greater absolute thromboembolic risk than CABG patients. The analysis compares adjusted relative hazards within each surgical population. Differences in baseline risk, patient characteristics, anticoagulation, follow-up duration, and outcome ascertainment prevent direct inference regarding absolute event rates across procedures.

### Relationship With Stroke-Specific Evidence

Stroke represents the most clinically consequential component of postoperative thromboembolic risk. Previous studies have demonstrated associations between cardiac surgery, POAF, and subsequent stroke [6-8]. However, stroke-specific and composite thromboembolic outcomes should not automatically be considered interchangeable.

Conen et al. evaluated new-onset perioperative AF following CABG and subsequent adverse outcomes in the CORONARY trial population [15]. Because the relevant long-term estimate was based on a stroke-specific endpoint, whereas the principal quantitative synthesis focused on sufficiently comparable broader thromboembolic outcomes, the Conen et al. estimate was not incorporated into the primary pooled analysis. It was instead considered as complementary stroke-specific evidence.

This distinction reduces endpoint mixing because composite thromboembolic outcomes may include ischemic stroke together with transient ischemic attack and/or systemic or peripheral embolic events, depending on the individual study. Pooling these outcomes indiscriminately with stroke-only endpoints could introduce additional clinical heterogeneity.

### Implications for Anticoagulation

The observed procedure-specific difference raises a clinically relevant question regarding whether thromboembolic risk assessment after POAF should account for the type of cardiac surgery. Current AF management guidance recognizes the importance of assessing thromboembolic risk and considering anticoagulation in appropriate patients [14]. However, management of POAF after cardiac surgery is more complex because potential thromboembolic benefit must be balanced against postoperative bleeding risk.

The present analysis did not evaluate the efficacy or safety of anticoagulation strategies and therefore does not establish that patients with POAF after valve surgery should routinely receive long-term anticoagulation or that anticoagulation strategies should differ between CABG and valve surgery. Rather, the findings generate the hypothesis that the prognostic significance of POAF may not be uniform across surgical populations.

Anticoagulation exposure also differed across observational cohorts and could change during follow-up. Consequently, differences in anticoagulation use may have influenced the observed associations and represent an important source of residual confounding.

### Comparison With Previous Evidence

Previous systematic reviews and meta-analyses have demonstrated associations between POAF and adverse outcomes following cardiac surgery [3,16]. Other studies have examined AF recurrence [9], long-term thromboembolic outcomes following CABG [5,6,13], and perioperative or long-term cardiovascular outcomes [15]. Collectively, this evidence suggests that POAF should not necessarily be regarded as a uniformly benign and self-limited postoperative event.

The present analysis extends this literature by focusing on procedure-stratified long-term thromboembolic associations and by formally comparing the pooled relative estimates for CABG and isolated valve surgery. Rather than treating cardiac surgery as a homogeneous exposure, this approach explores whether the prognostic association between POAF and subsequent thromboembolism may differ according to the index surgical procedure.

### Strengths

A principal strength of this analysis is the use of adjusted hazard ratios, rather than attempting to reconstruct unadjusted effects from incomplete event counts. The procedure-specific approach also reduces the clinical heterogeneity that would arise from combining CABG and valve surgery into a single cardiac-surgery population.

The primary analysis included 31,644 CABG patients [5,13] and 8,625 valve-surgery patients [17,18]. All four study estimates were directionally consistent, and the formal interaction analysis directly addressed the central hypothesis rather than inferring a difference from separate subgroup significance tests.The formal interaction analysis directly addressed the prespecified procedure-specific comparison rather than inferring effect modification from differences in statistical significance between the individual subgroups.

### Limitations

Several limitations should be considered when interpreting these findings. First, only four studies contributed to the primary quantitative synthesis, with two studies in each procedure subgroup. Estimates of between-study variance are therefore imprecise, and the observed I^2=0% in both subgroups should not be interpreted as evidence of complete methodological or clinical homogeneity.

Second, the analysis is based on observational associations and adjusted estimates reported by the original studies. Adjustment strategies differed among cohorts, and residual or unmeasured confounding cannot be excluded.

Third, definitions of thromboembolism were not completely uniform across the included studies. Differences in whether ischemic stroke, transient ischemic attack, systemic embolism, or peripheral arterial embolism contributed to the reported endpoint may have influenced the individual and pooled estimates.

Fourth, follow-up definitions and the handling of early postoperative events differed among studies, potentially affecting the distinction between genuinely long-term thromboembolic events and complications occurring during or shortly after the index hospitalization.

Fifth, anticoagulation practices were not standardized across studies. Differences in initiation, duration, adherence, and changes in anticoagulation during follow-up may have influenced the observed thromboembolic associations.

Sixth, the isolated valve-surgery subgroup remains clinically heterogeneous. The available evidence did not permit adequately powered separate meta-analyses according to individual valve procedure or underlying valve pathology.

Finally, with only two studies in each procedure subgroup, formal assessment of small-study effects or publication bias would not be meaningful. Although the between-subgroup interaction reached conventional statistical significance (P=0.043), this estimate is based on only four study-level effects and lies close to the conventional significance threshold. The interaction should therefore be regarded as exploratory and hypothesis-generating pending confirmation in additional independent cohorts.

### Clinical and Research Implications

The present findings support further investigation of procedure-specific risk following POAF rather than assuming that its long-term prognostic significance is uniform across all cardiac surgical populations. However, the current evidence is insufficient to support procedure-specific changes in anticoagulation or other long-term management strategies.

Future studies should prospectively distinguish CABG, aortic valve surgery, mitral valve surgery, and combined procedures rather than reporting POAF outcomes only for an aggregated cardiac-surgery population. Standardized definitions of POAF, recurrent AF, ischemic stroke, transient ischemic attack, systemic embolism, anticoagulation exposure, and follow-up time-zero would improve comparability across studies and strengthen future evidence synthesis.

Long-term rhythm surveillance may help determine whether recurrent or subclinical AF mediates the association between postoperative AF and subsequent thromboembolism, particularly given previous evidence demonstrating AF recurrence after hospital discharge [9].

Adequately powered prospective studies should also determine whether the apparent procedure-specific difference persists after standardized adjustment for baseline thromboembolic risk, anticoagulation exposure, and other clinically relevant confounders. Randomized evidence evaluating treatment strategies would ultimately be required before procedure-specific long-term anticoagulation recommendations could be justified.

## Conclusions

Postoperative atrial fibrillation after cardiac surgery may have important long-term thromboembolic implications, and its prognostic significance may differ according to the type of cardiac surgical procedure. These findings support considering the index surgical procedure when evaluating the longer-term clinical significance of POAF rather than treating POAF as a uniform postoperative phenomenon across all cardiac surgical populations.

The available evidence remains limited and does not establish that anticoagulation or other long-term management strategies should differ according to surgical procedure. Further prospective studies using standardized definitions of POAF, thromboembolic outcomes, recurrent atrial fibrillation, anticoagulation exposure, and follow-up are needed to clarify procedure-specific risk and determine whether such differences should influence postoperative surveillance or antithrombotic management.

## Data Availability

All data analyzed in this study were derived from previously published studies cited in the manuscript. The data used for the meta-analysis are contained within the manuscript. The study protocol was preregistered on OSF and is publicly available.

https://pubmed.ncbi.nlm.nih.gov

## Additional Information

### Author Contributions

All authors have reviewed the final version to be published and agreed to be accountable for all aspects of the work.

**Concept and design:** Asad Ullah

**Acquisition, analysis, or interpretation of data:** Asad Ullah

**Drafting of the manuscript:** Asad Ullah

**Critical review of the manuscript for important intellectual content:** Asad Ullah

**Supervision:** Asad Ullah

